# Assessing frailty in lung transplantation candidates

**DOI:** 10.64898/2026.09.24.26363958

**Authors:** B Planas-Pascual, I Bello, V Fernández-Piñar, ST Spiliopoulou, M Deu-Martín, G Ballesteros-Reviriego, C Berasetegui-García, M Ribas-Ball, A Gómez-Garrido

**Affiliations:** Rehabilitation and Physical Medicine Department Hospital Universitari Vall d’Hebron, Barcelona, Spain; Universitat de Vic – Universitat Central de Catalunya (UVic-UCC), Departament d’Atenció Clínica en Alteracions de Salut, Vic, Barcelona, Spain; Thoracic Surgeon Department. Hospital Universitari Vall d’Hebron, Barcelona, Spain; Pulmonologist Department. Hospital Universitari Vall d’Hebron, Barcelona, Spain; Anesthesiologist Department. Hospital Universitari Vall d’Hebron, Barcelona, Spain

**Keywords:** Lung transplantation, frailty, frailty assessment, SPPB, SHARE-FI, chronic obstructive pulmonary disease, COPD, interstitial lung disease (ILD)

## Abstract

**Background:** Frailty assessment is important while awaiting lung transplantation (LTx) as frailty is associated with disability and death before and after LTx. We assessed known risks factors and/or frailty indicators in frail/pre-frail/non-frail patients according to the ‘Short Physical Performance Battery’ (SPPB) and ‘Frailty Instrument for primary care of the Survey of Health, Ageing and Retirement in Europe’ (SHARE-FI) instruments. We also assessed the value of SHARE-FI as a proxy to detect frail patients.

**Methods:** Frailty was assessed using SPPB and SHARE-FI in 82 patients on a waiting list for LTx. Demographic, anthropomorphic, and clinical variables were collected. The patients underwent a respiratory/cardiorespiratory, biochemical, and quadriceps muscle performance assessment.

**Results:** The mean age was 57.3 ± 9.4 years. Chronic obstructive pulmonary disease (COPD) (41.5%) and interstitial lung disease (ILD) (30.6%) were the most common indications for LTx. Four (4.9%), 13 (15.9%), and 65 (79.3%) patients were classified as frail, pre-frail, and non-frail by SPPB. These figures were 26 (31.7%), 41 (50%), and 15 (18.3%) with SHARE-FI. SPPB found differences between frail/pre-frail patients in the physical performance (6MWD) and muscular assessment, while SHARE-FI found differences in age, 6MWD, and oxygen saturation, and a trend in muscular strength. For COPD and ILD, frailty discrimination was better with SPPB. Concordance between the two measurements was weak. All SPPB frail ILD patients were detected by SHARE-FI. *Conclusion.* In patients awaiting LTx, SPPB showed better discrimination than SHARE-FI for COPD and ILD. SHARE-FI seems appropriate as a preliminary screening instrument for patients with ILD.

## Introduction

Chronic respiratory diseases (CRDs) are associated with significant morbidity, disability, and premature mortality worldwide(1). Lung transplantation (LTx) is a therapeutic alternative for patients with advanced lung disease presenting with a high risk (>50%) of disease-related death and increased likelihood (>80%) of short and long-term survival. The proper selection of suitable candidates is of utmost importance in order to increase the chances of success(2). LTx has the potential to improve quality of life and prolong life expectancy(3), with a conditional median survival of 8.1 years in recipients who survive their first year post LTx(4).

Frailty is common in CRDs and is associated with poor functional status, disease severity, disability, poor health-related quality of life (HRQoL), and mortality(3, 5–7). This aspect is of special concern while awaiting LTx as frailty has been associated with disability and death before LTx(8, 9) and with a greater ‘vulnerability to stressors’(10); a deleterious relationship that worsens as waiting lists lengthen. Furthermore, some studies have found an association between preoperative frailty and increased 1-year mortality post LTx(11, 12). Frailty increases with age(13). In fact, aging-related signs that precede frailty, such as sarcopenia, are common in CRDs(3). Both the increasing age of recipients at the time of LTx observed in recent decades(2) and delays in performing the surgery, sometimes attributable to causes that are difficult to control, have again brought attention to frailty as a major consideration before LTx. Therefore, it is particularly important to identify frail patients while awaiting LTx as it allows us to implement interventions to optimize the patient’s situation before surgery(14).

Currently, there is no specific instrument for the assessment of frailty in patients awaiting LTx. The ‘Short Physical Performance Battery’ (SPPB) has been widely used in the assessment of frailty across generic populations (16), however its use in a pre LTx population is sporadic. In the Lung Transplant Body Composition study (8, 11) the SPPB was used as a frailty measure, demonstrating that frailty is associated with increased short-term mortality risk in patients awaiting LTx. Moreover, its value of containing functional tests such as standing balance and gait speed may help guide pre and post-operative rehabilitation programs in LTx populations. However, discussions around its complexity with regards to the physical space, special equipment and time required to complete the assessment, may affect its real-world practicality in a preoperative assessment setting (17). The ‘Frailty Instrument for primary care of the Survey of Health, Ageing and Retirement in Europe’ (SHARE-FI)(18) is another frailty metric that is intended to be used in community-dwelling adults aged 50 and over. It is a widely used tool due to its simplicity, however is based on a subjective assessment of a person’s disease-related limitations.

The objective of this study is to compare the SPPB and SHARE-FI instruments in the detection of frailty in patients awaiting LTx. It will assess the value of both assessment tools as a proxy to detect frail patients with the most common primary indications for LTx. This study is also the first to assess the use of the SHARE-FI in pre LTx frailty assessment.

## Materials and Methods

### Study subjects

This cross-sectional study was conducted at the Vall d’Hebron Hospital (Catalonia, Spain). Eighty-two patients on a LTx waiting list were included in this study. Patients were recruited between 1 July 2019 and 13 March 2020, when recruitment was halted due to the COVID-19 pandemic. The inclusion criteria for this study included adults between 18 and 70 years old, awaiting unilateral or bilateral LTx. All transplanted patients underwent an early post-operative rehabilitation program in addition to the standard rehabilitation regime provided by the physiotherapy service. Exclusion criteria for this study included any contraindication to participate in the rehabilitation program, such as pregnancy, pacemaker implantation or fractures requiring immobilization, as well as impaired cognitive function or psychiatric disorders. Patients who did not provide written informed consent to this study were also excluded. This study was approved by the Clinical Research Ethics Committee (CEIm) of Hospital Universitari Vall d’Hebron (approval number PR(AG)49/2019) and registered at ClinicalTrials.gov (NCT04244734).

### Variables

Demographic, anthropomorphic, and clinical variables were collected from medical records upon entering the study. These included age, sex, body mass index (BMI), primary indication for LTx, and the use of domiciliary oxygen therapy (DOT) and/or non-invasive mechanical ventilation (NMV). Severity of illness and need for transplant was included using the Lung Allocation Score (LAS) . The Charlson Comorbidity Index(19) was calculated on the basis of the available information. The following assessment took place upon entering the study: (i) cardiorespiratory tests, (ii) blood analyses, (iii) muscle strength. Cardiorespiratory tests included spirometry (forced expiratory volume 1 second (FEV1) and forced vital capacity (FVC)), maximal inspiratory pressure (MIP), maximal expiratory pressure (MEP), peak cough flow (PCF), oxygen saturation values, and 6-minute walking distance (6MWD). The lower limit of normal (LLN) values of MIP and MEP were calculated according to Evans and Whitelaw (20). Blood analyses included hemoglobin, total protein, albumin, aspartate aminotransferase (AST), alanine aminotransferase (ALT) and creatinine. Finally, muscle strength testing was done using quadriceps muscle performance (QMP) using dynamometer, whilst global muscle strength was measured by hand-grip dynamometry and the Medical Research Council Sum Score (MRC-SS). The hand-grip dynamometry used a cutoff value of <11kg for male and <7kg for female critically ill patients (21). The MRC-SS is an evaluation of six major muscle groups, where a score of less than 48 of 60 is indicative of significant weakness (23).

#### Frailty assessment

Frailty was assessed by the instruments SHARE-FI (18, 24, 25) and SPPB (16). SHARE-FI(18, 24) uses five constructs based on Fried’s frailty phenotype (18) selected by Santos-Eggimann et al. (26). A freely available online calculator individualized for both men and women allows individuals to be categorized as non-frail, pre-frail, or frail(18). For females, scores < 0.315 are considered non-frail, scores from 0.316 to 2.130 are pre-frail, and frail scores are values > 2.131. For males, scores < 1.211 indicate non-frail, scores from 1.212 to 3.005 are pre-frail, and scores > 3.006 are frail. The SPPB(16, 27) consists of three assessments scored on a 0-to 4-point scale, including a gait speed, chair stands and balance assessment. The final summary performance score ranges from 0 to 12, with higher scores indicating superior lower extremity function (i.e., less frailty). Patients with 0 – 6 points were classified as frail, with 7 – 9 points as pre-frail, and with 10 – 12 points as non-frail(28).

#### Statistical analysis

The “frail” and “pre-frail” SPPB frailty categories were combined with some of the relationships analyzed, as shown in the following tables, in order to strengthen the power of the analysis. Continuous variables are expressed as mean (standard deviation [SD])/median with 95% confidence interval (95% CI) and compared using the Kruskal Wallis test. Categorical variables are expressed as n (%) and compared using the Chi-square test or Fisher’s exact test. Cohen’s kappa coefficient (k) was calculated to assess the strength of the relationship between the SHARE-FI and SPPB frailty assessments in the most common primary indications for LTx. All the statistical analyses were performed using the “R” statistical package (R version 4.0.3. The R Foundation for Statistical Computing). The level of statistical significance was set at P < 0.05.

## Results

### Patients’ characteristics

The 82 patients included in this analysis had a mean (SD) age of 57.3 (9.4) years, 49 (59.8%) were male, with a mean BMI of 25.2 (4.3) kg/m^2^. Chronic obstructive pulmonary disease (COPD) was the most common indication for LTx (41.5%), followed by interstitial lung disease (ILD) (30.6%). The patients’ demographic, anthropometric, and clinical characteristics are shown in Table 1.

**Table 1.**
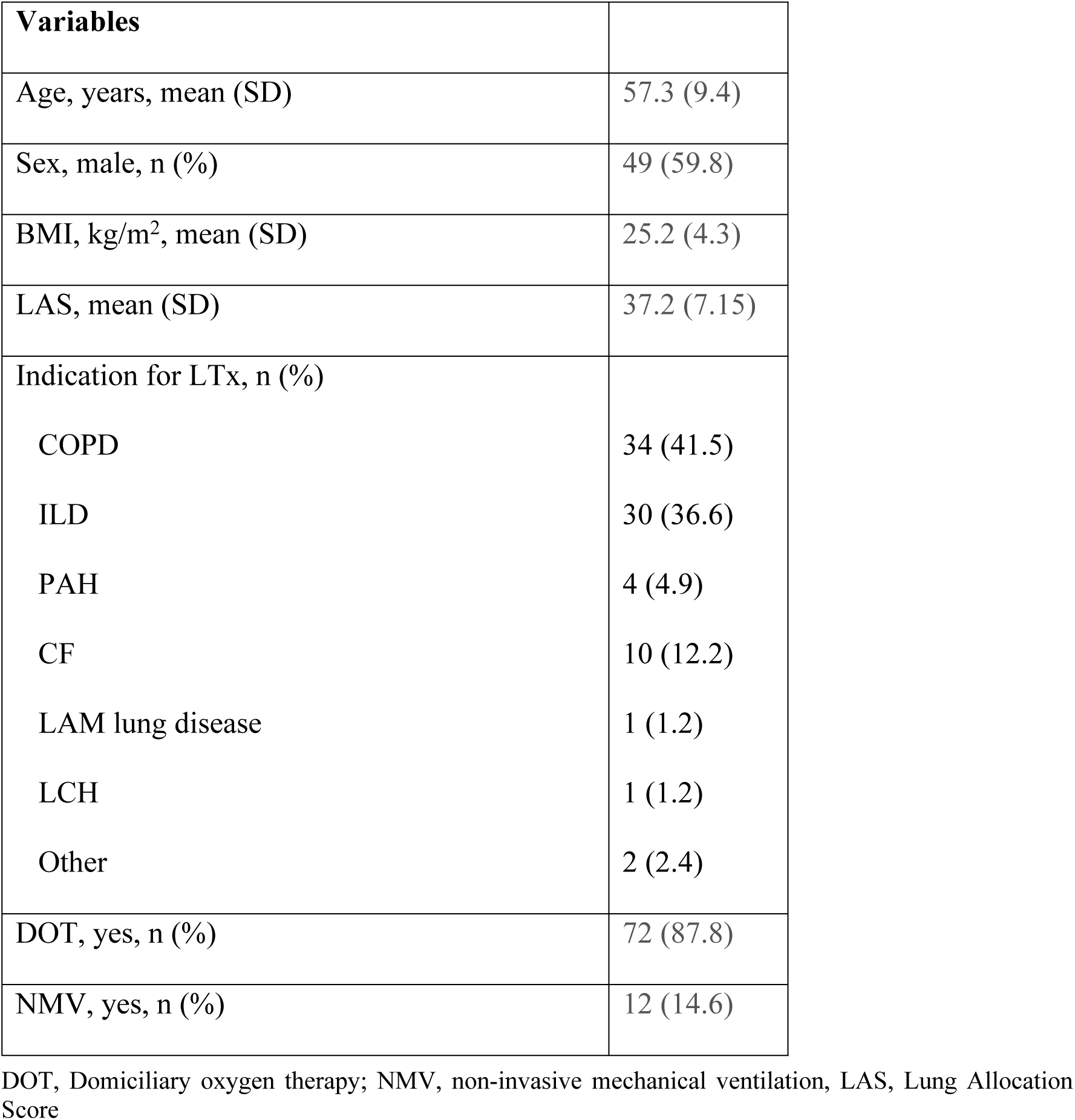
Demographic, anthropometric, and clinical characteristics of patients.

| Variables |  |
| --- | --- |
| Age, years, mean (SD) | 57.3 (9.4) |
| Sex, male, n (%) | 49 (59.8) |
| BMI, kg/m<sup>2</sup>, mean (SD) | 25.2 (4.3) |
| LAS, mean (SD) | 37.2 (7.15) |
| Indication for LTx, n (%) |  |
| COPD | 34 (41.5) |
| ILD | 30 (36.6) |
| PAH | 4 (4.9) |
| CF | 10 (12.2) |
| LAM lung disease | 1 (1.2) |
| LCH | 1 (1.2) |
| Other | 2 (2.4) |
| DOT, yes, n (%) | 72 (87.8) |
| NMV, yes, n (%) | 12 (14.6) |

DOT, Domiciliary oxygen therapy; NMV, non-invasive mechanical ventilation, LAS, Lung Allocation Score

The mean (SD) SPPB score was 10.8 (1.7), with 4 (4.9%) patients classified as frail, 13 (15.9%) as pre-frail, and 65 (79.3%) as non-frail. These figures were 26 (31.7%), 41 (50%), and 15 (18.3%) with SHARE-FI.

### Demographic, anthropometric, and clinical characteristics according to frailty categories

These are shown in Table 2. Frail and pre-frail patients were only significantly older than non-frail patients when these were classified with SHARE-FI (mean [SD] was 58 [11.2], 59.1 [7.3], and 51.4 [9.7] in frail, pre-frail, and non-frail patients, respectively; P = 0.014). Among the patients classified as frail/pre-frail by SPPB, COPD was the most common primary indication for LTx. Eleven out of 34 COPD patients (32.4%) vs. 4 out of 30 ILD patients (13.4%) were classified as frail/pre-frail by SPPB.

**Table 2.** Demographic, anthropometric and clinical characteristics according to frailty categories.

| Variable | SPPB frailty category |  | P-value | SHARE-FI frailty category |  |  | P-value |
| --- | --- | --- | --- | --- | --- | --- | --- |
|  | Frail +<br>Pre-frail<br>(n = 17) | Non-frail<br>(n = 65) |  | Frail<br>(n = 26) | Pre-frail<br>(n = 41) | Non-frail<br>(n = 15) |  |
| Age, years, mean (SD) | 60.5 (5.3) | 56.5 (10.1) | 0.259 | 58.0 (11.2) | 59.1 (7.3) | 51.4 (9.7) | **0.014** |
| Sex, male, n (%) | 9 (52.9) | 40 (61.5) | 0.715 | 15 (57.7) | 27 (65.9) | 7 (46.7) | 0.417 |
| BMI, kg/m<sup>2</sup>, mean (SD) | 24.4 (4.4) | 25.4 (4.3) | 0.366 | 25 (4.5) | 25.9 (4.1) | 23.8 (4.6) | 0.299 |
| Indication for LTx, n (%) |  |  | 0.302 |  |  |  | **0.395** |
| COPD | 11 (64.7) | 23 (35.4) |  | 9 (34.6) | 21 (51.2) | 4 (26.7) |  |
| ILD | 4 (7.7) | 26 (40.0) |  | 11 (42.3) | 13 (31.7) | 6 (40.0) |  |
| PAH | 0 (0) | 4 (6.2) |  | 2 (7.7) | 1 (2.4) | 1 (6.7) |  |
| CF | 1 (5.9) | 9 (13.8) |  | 3 (11.5) | 3 (7.3) | 4 (26.7) |  |
| LAM lung disease | 0 (0) | 1 (1.5) |  | 0 (0) | 1 (2.4) | 0 (0) |  |
| LCH | 0 (0) | 1 (1.5) |  | 1 (3.8) | 0 (0) | 0 (0) |  |
| Other | 1 (5.9) | 1 (1.5) |  | 0 (0) | 2 (4.9) | 0 (0) |  |
| Charlson score, mean (SD) | 3.2 (1.0) | 3.1 (1.0) | 0.953 | 3.2 (1.0) | 3.2 (1.0) | 2.7 (1.0) | 0.406 |

CF, cystic fibrosis; COPD, chronic obstructive pulmonary disease; ILD, interstitial lung disease; LAM, lymphangioleiomyomatosis; LCH, Langerhans cell histiocytosis; LTx, lung transplantation; NMV, non-invasive mechanical ventilation; PAH, pulmonary arterial hypertension; SD, standard deviation.

Among the patients classified as frail or pre-frail by SHARE-FI, ILD and COPD were the most common primary indications, respectively. Nine COPD patients (26.5%) vs. 11 ILD patients (36.7%) were classified as frail by SHARE-FI. These figures were 61.8% and 43.3% for pre-frail patients, respectively.

### Respiratory/cardiorespiratory tests

These are shown in Table 3. Frail/pre-frail patients showed lower 6MWD values when using SPPB. Frail patients showed the lowest 6MWD value when using SHARE-FI. Patients classified as frail/pre-frail only showed a lower FEV_1_ compared to non-frail patients with SPPB, the difference being quasi-significant (28.4 [19.6] vs. 37.7 [23.7] %, respectively; P = 0.055). Frailty was associated with reduced oxygen saturation when using SHARE-FI (84.5 [6.3], 89.2 [5.6], and 90.2 [4.0] % for frail, pre-frail, and non-frail patients, respectively; P = 0.001).

**Table 3.** Respiratory / Cardiorespiratory assessment according to frailty categories.

| Variable | SPPB frailty category |  | P-value | SHARE-FI frailty category |  |  | P-value |
| --- | --- | --- | --- | --- | --- | --- | --- |
|  | Frail + pre-frail<br>(n = 17) | Non-frail<br>(n = 65) |  | Frail<br>(n = 26) | Pre-frail<br>(n = 41) | Non-frail<br>(n = 15) |  |
| FEV<sub>1</sub>, % predicted | 28.4 (19.6) | 37.7 (23.7) | 0.055 | 35 (20.8) | 33.6 (22.9) | 43.1 (27.3) | 0.377 |
| FVC, % predicted | 42.5 (17.3) | 48.8 (20.2) | 0.181 | 43.6 (17.2) | 47.4 (18.4) | 54.3 (25.8) | 0.446 |
| MIP, cmH<sub>2</sub>O | 61.6 (31.8) | 74.9 (30.1) | 0.104 | 67 (33.5) | 71.1 (31.2) | 83.7 (22.0) | 0.122 |
| LLN | 43.0 (11.1) | 46.5 (9.3) | 0.333 | 45.3 (10.3) | 46.4 (9.3) | 44.6 (10.3) | 0.664 |
| MEP, cmH<sub>2</sub>O | 95.2 (34.5) | 107.6 (34.7) | 0.094 | 100.8 (35.2) | 106.7 (36.3) | 107.9 (31.7) | 0.646 |
| LLN | 63.8 (4.9) | 66.0 (6.8) | 0.175 | 65.6 (7.0) | 65.4 (5.4) | 69.8 (7.8) | 0.145 |
| PCF, L/min | 245.3 (135.6) | 318.3 (165.1) | 0.103 | 324.2 (148.5) | 283.1 (167.6) | 321.3 (169.4) | 0.391 |
| Oxygen saturation, % | 87.3 (7.0) | 88.0 (5.8) | 0.706 | 84.5 (6.3) | 89.2 (5.6) | 90.2 (4.0) | **0.001** |
| 6MWD, meters | 193.2 (85.1) | 315 (121.8) | **<0.001** | 223.9 (105.8) | 297.5 (112.3) | 383.0 (129.8) | **<0.001** |

All values are expressed as mean (SD)
FEV<sub>1</sub>, forced expiratory volume in 1 second; FVC, forced vital capacity; LLN, lower limit of normal; MEP, maximal expiratory pressure; MIP, maximal inspiratory pressure;
PCF, peak cough flow; SD, standard deviation; 6MWD, 6-minute walking distance.

### Biochemical values

These are shown in Table 4. No differences between frailty categories were observed in hemoglobin, total protein, albumin, AST, ALT, or creatinine plasma values for either of the frailty assessment instruments used.

**Table 4.** Biochemical values according to frailty categories.

| Variable | SPPB frailty category |  | P-value | SHARE-FI frailty category |  |  | P-value |
| --- | --- | --- | --- | --- | --- | --- | --- |
|  | Frail + pre-frail<br>(n = 17) | Non-frail<br>(n = 65) |  | Frail<br>(n = 26) | Pre-frail<br>(n = 41) | Non-frail<br>(n = 15) |  |
| Hemoglobin, g/dL | 14.1 (1.6) | 14.4 (1.8) | 0.651 | 14.4 (1.8) | 14.5 (1.5) | 13.6 (2.3) | 0.170 |
| Total protein, g/dL | 6.8 (0.6) | 7.1 (0.6) | 0.087 | 7.1 (0.6) | 7.0 (0.5) | 7.0 (0.8) | 0.833 |
| Albumin, g/dL | 4.2 (0.4) | 4.1 (0.4) | 0.666 | 4 (0.5) | 4.2 (0.3) | 4.0 (0.4) | 0.086 |
| AST, U/L | 22.2 (6.5) | 23.3 (7.3) | 0.840 | 24.4 (6.8) | 22.7 (7.3) | 21.6 (6.9) | 0.395 |
| ALT, U/L | 23.0 (9.9) | 22.9 (10.1) | 0.991 | 22.3 (9.6) | 24.6 (10.9) | 19.3 (7.2) | 0.264 |
| Creatinine, mg/dL | 0.8 (0.3) | 0.7 (0.2) | 0.689 | 0.8 (0.3) | 0.7 (0.2) | 0.8 (0.2) | 0.994 |

All values are expressed as mean (SD).
ALT, alanine aminotransferase; AST, aspartate aminotransferase; SD, standard deviation.

### Muscular strength/endurance

These are shown in Table 5. The population of frail/pre-frail patients according to SPPB showed a reduced MRC-SS (mean [SD] was 55.7 [4.4] vs. 58.3 [2.5] in non-frail patients; P = 0.008) and right and left QMP (15.6 [6.0] vs. 20.6 [6.0] kg, P = 0.004 and 14.6; [5.4] vs. 20.1 [6.5] kg, P = 0.002, respectively). No differences were observed when using SHARE-FI.

**Table 5.** Muscular assessment according to frailty categories.

| Variable | SPPB frailty category |  | P-value | SHARE-FI frailty category |  |  | P-value |
| --- | --- | --- | --- | --- | --- | --- | --- |
|  | Frail + pre-frail<br>(n = 17) | Non-frail<br>(n = 65) |  | Frail<br>(n = 26) | Pre-frail<br>(n = 41) | Non-frail<br>(n = 15) |  |
| Hand-grip strength, kg | 29.0 (8.4) | 32.2 (8.2) | 0.208 | 29.3 (8.2) | 31.8 (6.9) | 34.8 (11.0) | 0.213 |
| MRC-SS | 55.7 (4.4) | 58.3 (2.5) | **0.008** | 57.4 (3.4) | 57.9 (2.7) | 58.0 (4.0) | 0.387 |
| QMP (right), kg | 15.6 (6.0) | 20.6 (6.0) | **0.004** | 19.6 (6.6) | 18.6 (5.0) | 22.4 (8.4) | 0.208 |
| QMP (left), kg | 14.6 (5.4) | 20.1 (6.5) | **0.002** | 19.1 (7.4) | 17.5 (5) | 22.8 (8.2) | 0.077 |

All values are expressed as mean (SD)
MRC-SS, Medical Research Council sum score; QMP, quadriceps muscle performance; SD, standard deviation.

### Frailty concordance of SHARE-FI and SPPB for COPD and ILD patients

Of the nine frail COPD patients according to SHARE-FI, four were pre-frail and five were non-frail according to SPPB. Of the eleven frail ILD patients, three were frail and one was pre-frail by SPPB (all frail and pre-frail patients), and seven were non-frail according to SPPB. Cohen’s kappa coefficient for the concordance of frailty assessments was -0.02 (P = 0.786) for COPD patients and 0.097 (P = 0.109) for ILD patients.

## Discussion

This is the first study that compares the SHARE-FI and SPPB assessment tools in the detection of frailty in a pre-LTx population. In summary, our results confirm that COPD and ILD are the most common primary indications for LTx. The SPPB found differences between frail/pre-frail and non-frail patients in the physical performance (6MWD) and muscular assessment. Besides differences in 6MWD, SHARE-FI also found differences in age and oxygen saturation. Frailty discrimination was better with SPPB for the main primary indications for LTx. The concordance between the two measures was weak, although SHARE-FI seems to have a role as a proxy for frailty detection in patients with ILD.

The SPPB has been developed as a screening instrument to detect frailty syndrome in community-dwelling older adults and has demonstrated its value in the pre-LTx setting(8, 11). Four out of 82 patients awaiting LTx in our hospital were classified by SPPB as frail and 13 as pre-frail. In accordance with its objective nature and physical approach, we found differences in the physical performance and muscular assessment (MRC-SS and overall QMP [right and left]), which were significantly lower in frail/pre-frail patients vs. non-frail patients. This can be expected since reduced physical function and low muscle strength are included in the diagnostic criteria of sarcopenia(29). On the contrary, no differences were found in known risk factors for LTx prognosis such as age, sex, life expectancy (comorbidity index), lung function, or biochemical values.

The SHARE-FI, which is intended to facilitate the adoption of the frailty paradigm in primary care(18, 24, 25), has –to our knowledge-never been used to assess frailty in patients awaiting LTx. Unlike the SPPB, a main limitation of the SHARE-FI is its subjectivity where four of its five items may give rise to sex, age and cultural biases regarding self-perceived limitations in activities of daily living. This instrument classified 26 patients as frail and 41 as pre-frail. Nevertheless, this instrument did find significant differences in physical performance (6MWD). According to SHARE-FI, frail patients were also significantly older, which points to a worse self-perception in the elderly, as mentioned previously. However, no differences in self-perception were observed between men and women, which may be explained by differences in the interpretation of results for both sexes in this instrument. We found a difference in the percentage of oxygen saturation, which was lower in frail patients than in pre-frail and non-frail patients. This result may explain the poor physical status expressed by these patients, which was not captured by SPPB despite it being objective. This relationship should be better explored. The only objective measurement in the SHARE-FI is muscle hand-grip strength, which is not included in SPPB. Although a trend towards a lower hand-grip strength was observed, this difference was not statistically significant. A similar trend was observed for QMP (right and left) but not for MRC-SS.

Given the ease of use of SHARE-FI in settings with limited time per patient such as primary care, we also assessed its value as a preliminary screening instrument to detect frail patients so that more accurate screening instruments such as the SPPB could be administered subsequently to a selected group of patients instead of indiscriminately. Given the differences in clinical manifestations of the different primary indications for LTx, how they evolve, and how they affect activities of daily living (ADLs), we only focused on the two most common ones: COPD and ILD. None of the 34 patients with COPD were classified as frail by SPPB, while nine were classified as frail by SHARE-FI. Four of these were pre-frail according to SPPB, which leaves seven pre-frail patients unaccounted for. These are likely to be included in the pre-frail category of SHARE-FI; however, given that only four patients were non-frail according to this instrument, this first screening did not explain this. We observed greater self-perceived frailty (understood as a worse self-perception of the ability to perform ADLs) in this disease with respect to the objective SPPB measurement. Although the onset of symptoms in COPD and the evolution of the disease are slow and patients are likely to get used to their quality of life, the relative youth of the patients included in this study and their poor health status may explain this result. Despite the low concordance of both measurements in ILD patients, four of the thirty patients with this disease were classified as frail by SPPB, while eleven were classified as frail by SHARE-FI. Three of these were frail and one was pre-frail according to SPPB, which covers all the frail and pre-frail ILD patients. Unlike in COPD, the onset of symptoms in ILD is insidious and the evolution of the disease is faster, resulting in the subjective and objective measurements of the effect of the disease being similar, at least in our population. Finally, the inclusion of cystic fibrosis may have provided a skewed reality of the average age of our study populations, as this is a sub-population where LTx may be considered as a treatment option at a younger age.

The main limitation of this study is the generalisability of its findings across a worldwide population of people undergoing pre-LTx assessment. Moreover, our finding of better frailty detection in COPD and ILD illnesses using the SPPB may be argued as the sample sizes available in PAH and CF are small. Therefore, we cannot be sure of the degree to which these measures can discriminate frailty in other respiratory diagnoses. This may be a relationship that should be explored further in future studies with larger sample sizes.

To summarize, in patients awaiting LTx, SPPB showed better discrimination than SHARE-FI for the most common primary indications for LTx. Both instruments detected differences in physical performance between patients with any degree of frailty and non-frail patients. SPPB also detected differences in muscular endurance, while SHARE-FI detected differences in age and oxygen saturation. SHARE-FI seems appropriate as a preliminary screening instrument for patients with ILD. The results of the study have to be interpreted with caution, especially given the small number of patients included.

## Author contributions

BPP and AGG take responsibility for the content of the manuscript, including the data analysis. Design (BPP and AGG), data collection (BPP, GBR, VF, and AGG). Interpretation of data (BPP, IB, AGG), drafting and revising of work (all authors), final approval (all authors).

## Data Availability

The data underlying this study contain potentially identifying and sensitive patient information and cannot be made publicly available due to restrictions imposed by the Clinical Research Ethics Committee (CEIm) of Hospital Universitari Vall d'Hebron (approval PR(AG)49/2019) and the terms of participants' informed consent, which did not include provisions for public data sharing. De-identified data are available upon reasonable request to qualified researchers who meet the criteria for access to confidential data, subject to approval by the Vall d'Hebron Hospital Research Institute (VHIR). Requests may be directed to the corresponding author.

## Acknowledgments

The authors would like to thank Beatriz Viejo, PhD for her assistance in writing and editing this manuscript and the Statistics and Bioinformatics Unit (UEB) of Vall d’Hebron Hospital Research Institute (VHIR) for performing the statistical analysis.

## List of non-standard abbreviations

N/A

